# Computational network models for forecasting and control of mental health trajectories in digital applications

**DOI:** 10.1101/2025.07.03.25330825

**Authors:** Janik Fechtelpeter, Christian Rauschenberg, Christian Goetzl, Selina Hiller, Niklas Emonds, Silvia Krumm, Ulrich Reininghaus, Daniel Durstewitz, Georgia Koppe

## Abstract

Ecological momentary assessments (EMA) have transformed mobile mental health by capturing real-time fluctuations in psychological states and behavior. While forecasting future states from EMA data is crucial for adaptive interventions, most current approaches to modeling the underlying psychological mechanisms rely on linear assumptions. These include common network based methods such as vector autoregression (VAR) or Kalman filtering, which assume fixed and proportional relationships among variables. However, a growing body of evidence suggests that psychological dynamics exhibit nonlinear properties raising concerns about the adequacy of linear models for both interpretation and prediction.

Here, we leverage three independent 40-day micro-randomized trials (N=145) to benchmark a spectrum of models—from naïve baselines and linear network models to autoregressive Transformers and nonlinear state-space models (SSMs) built on piecewise-linear recurrent neural networks (PLRNNs). PLRNNs provided the most accurate forecasts, including predictions of how individuals responded to interventions. Beyond superior forecasting, the PLRNN’s latent-network structure allowed us to simulate how changes in individual psychological states spread through the system. This revealed interpretable patterns of influence—highlighting central network nodes like *sad* or *down* as high-impact intervention targets based on their strong ripple effects. Critically, performance remained robust under real-time retraining constraints and varying data completeness, underscoring the practical viability of nonlinear SSMs in deployed mobile mental health systems. Our results establish PLRNN-based forecasting as a powerful, interpretable foundation for real-time, model-predictive control of digital mental health.

## 1 Introduction

Digital mental health interventions, mobile mental health (mHealth) applications in particular, provide a cost-effective and accessible way for individuals with mental health conditions to access evidence-based mental health services [1, 2], bridge waiting periods before psychotherapy [3, 4], or supplement ongoing treatment in blended care models [5]. These apps increasingly incorporate Ecological Momentary Assessments (EMA) to capture real-time psychological states and behavior, enabling the continuous tracking of mental health in daily life to inform the delivery of intervention components that are more tailored to moment, context, and person, including Ecological Momentary Interventions (EMIs) or Just-in-time adaptive interventions (JITAIs) [6–10]. EMA data - such as self-reported mood, anxiety, stress, worry, or arousal levels - are essential for identifying critical moments when interventions may be most effective, such as detecting early signs of depression or escalating anxiety that warrant timely coping strategies [7, 11, 12].

A growing body of research suggests that EMA variables are not isolated indicators but interact within complex psychological networks [13, 14]. This network approach aligns closely with the perspective that psychological functioning is best understood as a dynamical system (DS) [15, 16], where individual states influence each other across multiple time scales [17–20]. For example, network models have been employed to elucidate how therapy attenuates the mutual interactions between anxiety and avoidance [21], how transdiagnostic irritability is driven by frustration [22], how more generally emotional states can shift nonlinearly between steady and oscillatory patterns [23], and how the interactions between internal and external factors can give rise to psychiatric symptoms [24]. Moreover, they offer insights into how EMI can promote adaptive change over time [25]. These insights are increasingly being used to tailor mHealth interventions to individual needs, enhancing personalization and clinical relevance [11, 25–28].

Most recent research uses linear methods such as vector autoregressive models [29], hierarchical linear modeling [30, 31], and mixed-effects models [32], to analyze EMA data. These methods are widely used because they offer interpretability and simplicity in modeling temporal relationships between psychological variables.

However, psychological time series often show clear signatures of nonlinear dynamics, including multistability [33] and phase transitions linked to sudden mood shifts, depressive episode onset, and recovery trajectories [34]. Linear models on the other hand assume fixed, proportional relationships between variables [35] and are fundamentally limited in their ability to capture such dynamics. This raises concerns about their suitability for forecasting psychological states, especially in contexts where accurate, real-time prediction is essential for adaptive intervention, such as in model-predictive control frameworks.

Here, using three independent and comparable datasets, we robustly demonstrate that forecasting models accounting for nonlinear interactions between dynamic variables significantly outperform linear approaches in predicting psychological states, providing a more valid and realistic generative framework. In addition to improved predictive accuracy, these non-linear models yield more interpretable network structures, shedding light on the underlying dynamics of psychological state interactions. Specifically, we show that certain classes of nonlinear dynamical models not only illuminate the architecture of psychological networks but also help to identify the most promising targets for intervention. Taken together, these findings highlight the critical importance of nonlinear modeling for guiding the personalized delivery of EMIs. The modeling results are highly replicable and generalize to real-time deployment.

## 2 Results

### Forecasting Psychological States from EMA and EMI Data

While EMAs enable real-time assessment and monitoring of psychological states and timely interventions in mHealth applications including EMI and JITAI, the predominant use of linear models has constrained their ability to capture the complex, nonlinear interactions inherent to psychological processes. To overcome these limitations, we systematically evaluated the predictive accuracy and interpretability of various forecasting models using three unique EMA+EMI datasets collected over 40 days, with a total sample size of *N* = 145, in the AI4U living lab [36]. More details on the project can be found in [37, 38]. More specifically, we employed an online training/evaluation scheme which imitates how such models would be deployed in a real-world scenario (see Fig. 1A). We compared a set of model classes ranging from simple, static baselines to advanced dynamic network models, including first order vector autoregressive models (VAR(1)), linear state-space models (Kalman filters), nonlinear state-space models (SSMs) incorporating latent piecewise-linear recurrent neural networks (PLRNN) [39–41], and autoregressive Transformer architectures [42, 43] (see Appx. Methods A.5 for rationale behind these choices). Hyperparameter tuning was conducted exclusively on the first dataset (sample 1, see Appx. A.6), while sample 2 and sample 3 served as independent test sets (see Methods A.3 for details).

**Figure 1.**
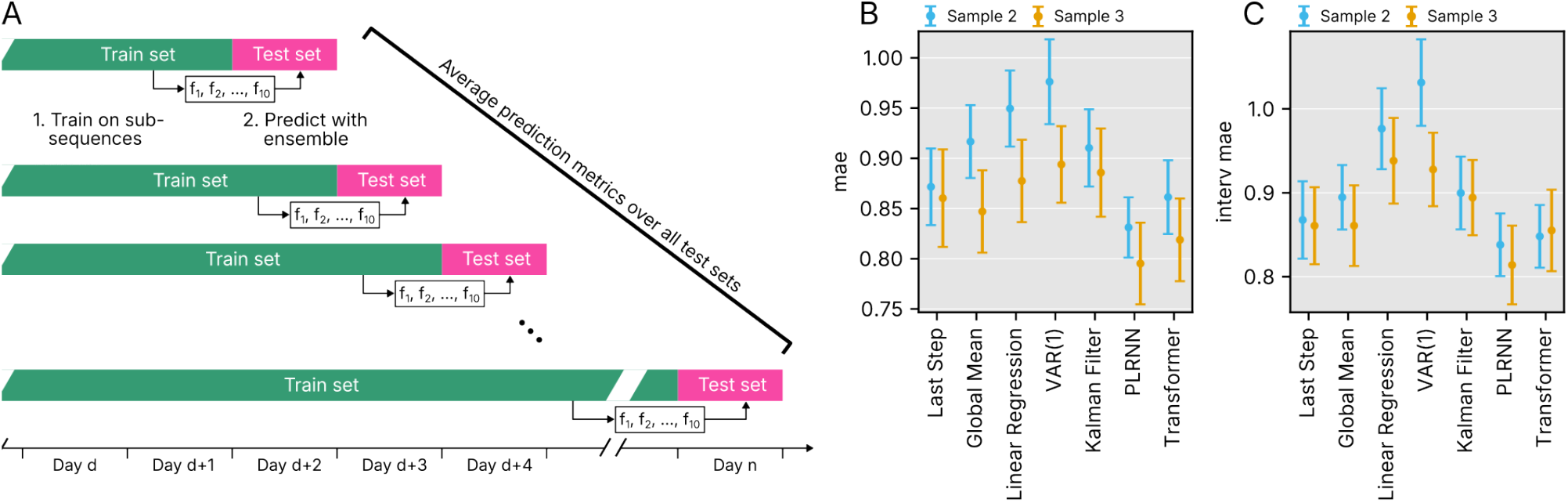
A: Online model training/evaluation procedure. For each day *d*, the train set included the entire time series up to (but not including) day *d*, while the test set consisted of data from day *d* itself. For each train set, 10 models were trained with independent random initializations, and their average prediction was used as the forecast. Evaluation metrics were computed across all test days. B: Mean absolute error (MAE) for predicted absolute EMA values across all tested models. C: MAE of time points immediately succeeding an EMI.

### Nonlinear Models Significantly Outperform Linear Approaches

Across the two independent replication samples (sample 2 with 48 and sample 3 with 51 participants, respectively), nonlinear forecasting models consistently outperformed linear methods. Specifically, the PLRNN model significantly improved prediction accuracy of daily psychological state fluctuations, achieving lower mean absolute errors (MAE; sample 2 = 0.831, sample 3 = 0.795, Fig. 1B) compared to VAR models in both samples, and compared to Kalman filter models in sample 2. Detailed results can be fount in Table 1. Notably, linear models even performed significantly worse than simple baseline approaches, such as using the previous time step or the overall expectation as predictors. Adding to that, PLRNN models also outperformed linear models in predicting EMA states immediately following an EMI (sample 2 = 0.838, sample 3 = 0.814, Fig. 1C), highlighting its potential for informing interventions.

**Table 1.** Evaluation metrics for all tested models in samples 2 and 3. Mean and standard error are displayed. Additional paired t-test results for the comparison of the absolute score MAE between each model and the best performing model (PLRNN), with Bonferroni-corrected *p* values. Lower degrees of freedom in the Kalman filter are a result of non-convergence for a few subjects.

| Sample | Model | MAE | Interv. MAE | MAE compared to PLRNN |
| --- | --- | --- | --- | --- |
| 2 | Last step | 0.872 ± 0.0382 | 0.868 ± 0.0461 | $t(47) = 2.14$, $p_{\text{Bonf.}} = .228$ |
| | Global Mean | 0.917 ± 0.0363 | 0.895 ± 0.0384 | $t(47) = 5.71$, $p_{\text{Bonf.}} < .001$ |
| | Linear Regression | 0.950 ± 0.0379 | 0.976 ± 0.0482 | $t(47) = 8.44$, $p_{\text{Bonf.}} < .001$ |
| | VAR(1) | 0.976 ± 0.0422 | 1.031 ± 0.0515 | $t(47) = 4.81$, $p_{\text{Bonf.}} < .001$ |
| | Kalman filter | 0.910 ± 0.0385 | 0.900 ± 0.0434 | $t(45) = 3.86$, $p_{\text{Bonf.}} = .002$ |
|  | PLRNN | **0.831 ± 0.0300** | **0.838 ± 0.0374** | — |
| | Transformer | 0.861 ± 0.0367 | 0.848 ± 0.0374 | $t(47) = 1.88$, $p_{\text{Bonf.}} = .401$ |
| 3 | Last step | 0.860 ± 0.0485 | 0.861 ± 0.0459 | $t(50) = 2.59$, $p_{\text{Bonf.}} = .075$ |
| | Global Mean | 0.847 ± 0.0410 | 0.861 ± 0.0481 | $t(50) = 6.56$, $p_{\text{Bonf.}} < .001$ |
| | Linear Regression | 0.877 ± 0.0410 | 0.938 ± 0.0511 | $t(50) = 8.31$, $p_{\text{Bonf.}} < .001$ |
| | VAR(1) | 0.894 ± 0.0381 | 0.928 ± 0.0438 | $t(50) = 5.59$, $p_{\text{Bonf.}} < .001$ |
| | Kalman filter | 0.886 ± 0.0439 | 0.894 ± 0.0449 | $t(45) = 2.26$, $p_{\text{Bonf.}} = .172$ |
|  | PLRNN | **0.795 ± 0.0407** | **0.814 ± 0.0469** | — |
| | Transformer | 0.819 ± 0.0411 | 0.855 ± 0.0485 | $t(50) = 2.75$, $p_{\text{Bonf.}} = .050$ |

### Enhanced Interpretability of Nonlinear Network Models

Beyond predictive performance, the PLRNN also markedly enhanced interpretability, which was uniquely facilitated by the specific architecture of this model. Because the PLRNN model incorporates a piecewise-linear activation structure, local derivatives can be analytically derived (see Methods 4). This capability enabled us to transparently reconstruct interpretable psychological network dynamics directly from the model. Fig. 2A,B depicts the effective connectivity of the PLRNNs in observation space, delivering directly interpretable behavioral contingencies, consistent across the sample, Bonferroni corrected for multiple comparisons.

**Figure 2.**
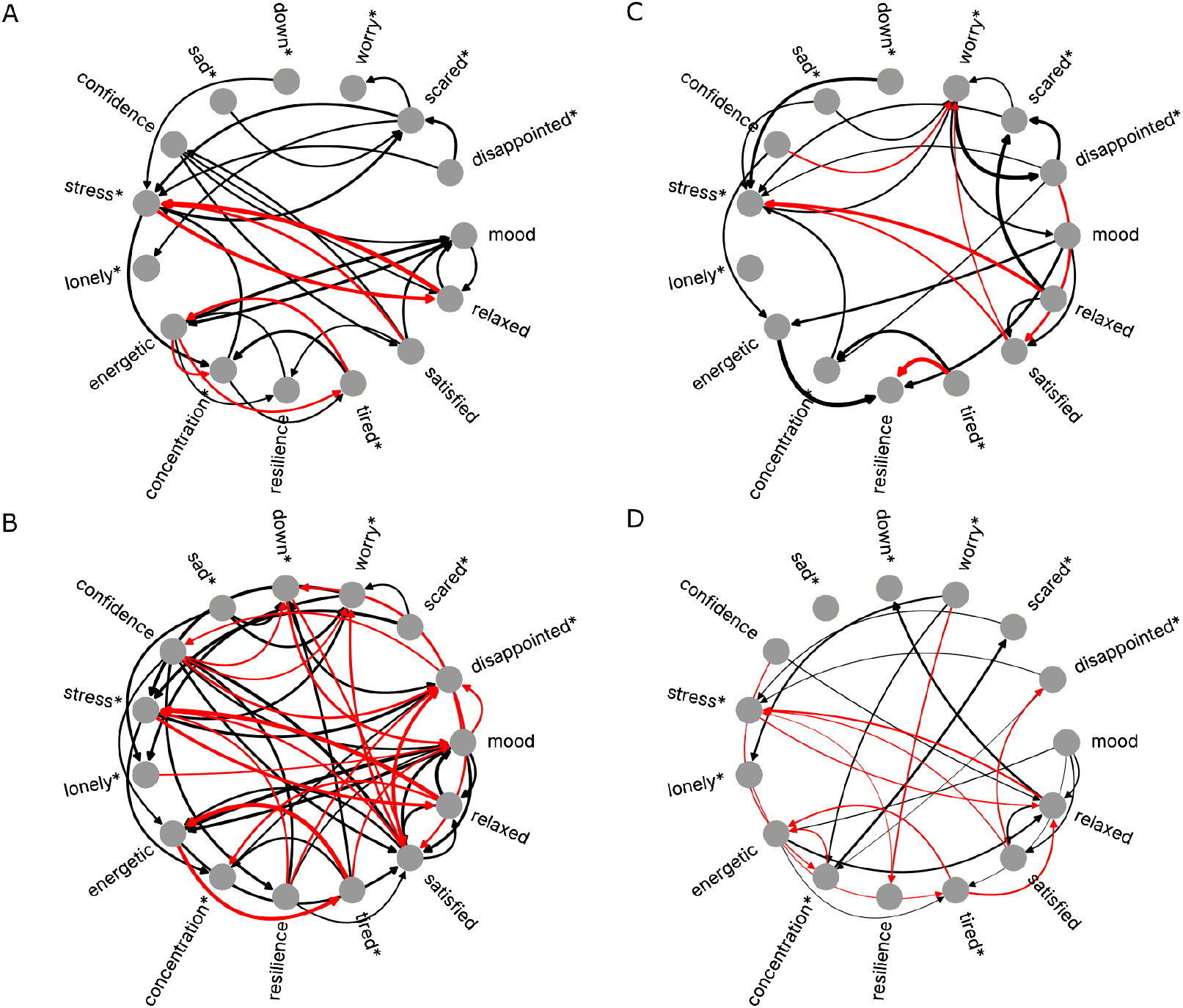
Models depicted as ideographic networks. Arrow width depicts mean edge strength over all participants. Positive connections are shown in black, negative connections in red. For visualization purposes, negatively poled items were not recoded and marked with an asterisk. A: PLRNN in sample 2 (Bonferroni corrected for multiple comparisons). B: PLRNN in sample 3 (Bonferroni corrected). C: VAR(1) in sample 2 (uncorrected). D: VAR(1) in sample 3 (uncorrected).

Across all participants, several EMA items exhibit significant connectivity - for example, *stress* and *relaxed* are mutually inhibitory, so are *tired* and *energetic*\*. In contrast, *mood* and *relaxed*, as well as *confidence* and *satisfied* are mutually excitatory. While significance levels differed between samples, the overall connectivity pattern remained largely consistent across samples and was intuitively interpretable. In contrast, connectivity inferred from VAR(1) models was notably more sparse (in fact, no connections survived multiple test corrections) and occasionally counterintuitive (see Fig. 2C, D; note that the VAR(1) networks were left uncorrected since nearly no connections would have survived multiple testing). For example, according to VAR(1), being *energetic* reinforced being *tired*, and being *relaxed* increased the likelihood of feeling *scared*. Such findings may stem from a fundamental limitation of traditional VAR modeling, as it cannot account for dynamics arising from moderating latent variables. Instead, VAR models must attribute all dynamics directly to observed variables, which may compromise interpretative validity. Networks inferred from Kalman filter models exhibited almost no significant connections, even without Bonferroni correction, underlining that the consistency of PLRNN networks is both due to nonlinearity and being an SSM. Moreover, although Transformer architectures achieved reasonable forecasting performance, they cannot be interpreted as ideographic networks - analogous to many other deep neural networks - and require longer training times, making them less suitable for real-time deployment in e.g. digital mental health interventions (see Appx. Fig. B1).

### Identifying Targets for Personalized Intervention

Exploiting the PLRNN’s explicit latent-network formulation — which admits analytical perturbations of individual nodes (see Methods 4 and Appx. Methods A.8) — we next examined how targeted disturbances of single psychological states ripple through the network to shape overall mental-health trajectories. Our analysis of the network dynamics revealed that certain states exert a disproportionately strong influence. In particular, nodes such as “down” and “sad” exhibited high relative cumulative impulse responses (rCIRs), marking them as promising targets for intervention (Fig. 3B). In contrast, other states—such as “tired”—showed weaker ripple effects, suggesting they may be less effective. The computed rCIR metric also carried an intuitively interpretable meaning in terms of the network’s structure: states with high rCIRs tended to occupy more central positions in the psychological network, exerting influence over multiple interconnected nodes (Spearman’s *ρ* = 0.832*/*0.836 in sample 2/3, *p <* .001, see Fig. 3C). When directed to the nodes with lowest out centrality, external inputs induced a significantly smaller absolute rCIR than when directed to the nodes with highest weighted out-degree centrality (*t*(114) = −4.48, *p <* .001), indicating the importance of strategically selecting intervention targets based on network centrality and rCIRs. However, rCIR has the advantage of quantifying predicted effects on a single-item resolution, while centrality only provides a system-wide measure. Crucially, the ability to specifically target isolated EMA nodes for mechanistic insight stems from the PLRNN’s unique architecture, enabling analytically tractable perturbation analyses. These findings provide an evidence-based framework for tailoring interventions to individual psychological network structures, that may optimize the effectiveness of EMIs, and help identifying candidate intervention targets.

**Figure 3.**
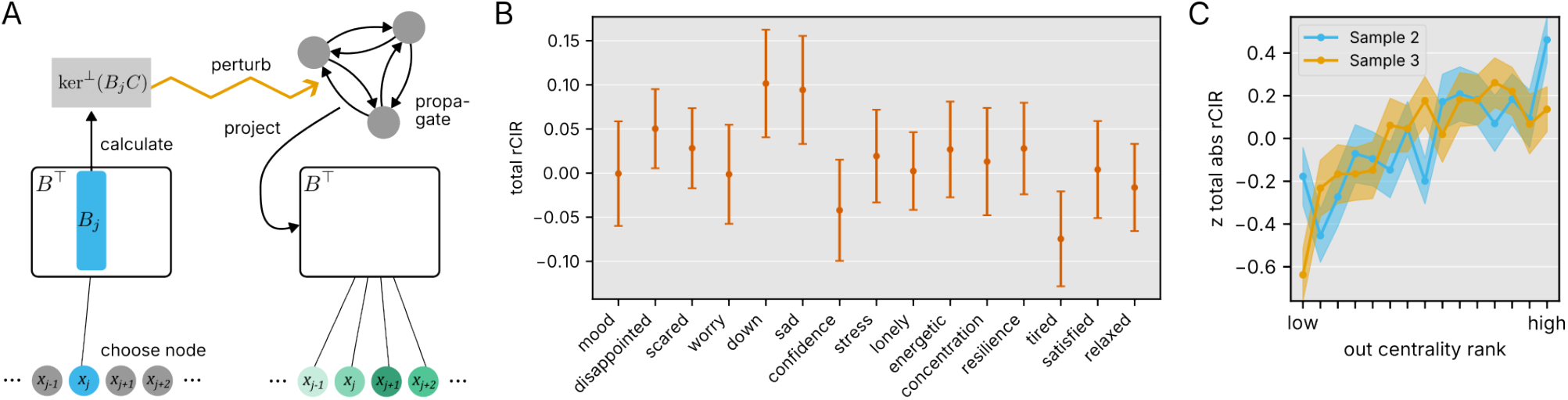
A: Perturbation procedure for latent models. External inputs are added to the latent time series. To perturb a certain observation node *j*, we compute the orthogonal complement to the kernel of row *j* of observation matrix *B* and use linear combinations of its elements as external inputs on the latent model. To compute the CIR_*T*_, the latent model is iterated *T* time steps, and the resulting predictions are summed. B: Total rCIR to inputs targeting each individual EMA item. Mean and standard error are displayed. C: Total rCIR (standardized within subjects) to inputs targeting specific nodes, plotted against the out-degree centrality rank of these nodes (higher rank number=higher centrality). The solid line indicates mean rCIR, shaded areas the standard error of the mean.

We also applied this novel approach to forecast and evaluate the effects of the administered EMIs by perturbing the inferred networks and computing their rCIR trajectories over a 24-hour period (see Appx. Methods A.8). Although direct validation is challenging due to the influence of multiple uncontrolled external factors in the empirical EMA time series, the model again produced plausible predictions: most EMIs were associated with either significantly positive or neutral effects on mental health (see Fig. 4C, D). Only one EMI was linked to adverse predicted outcomes; notably though, this pattern was consistent across both samples.

**Figure 4.**
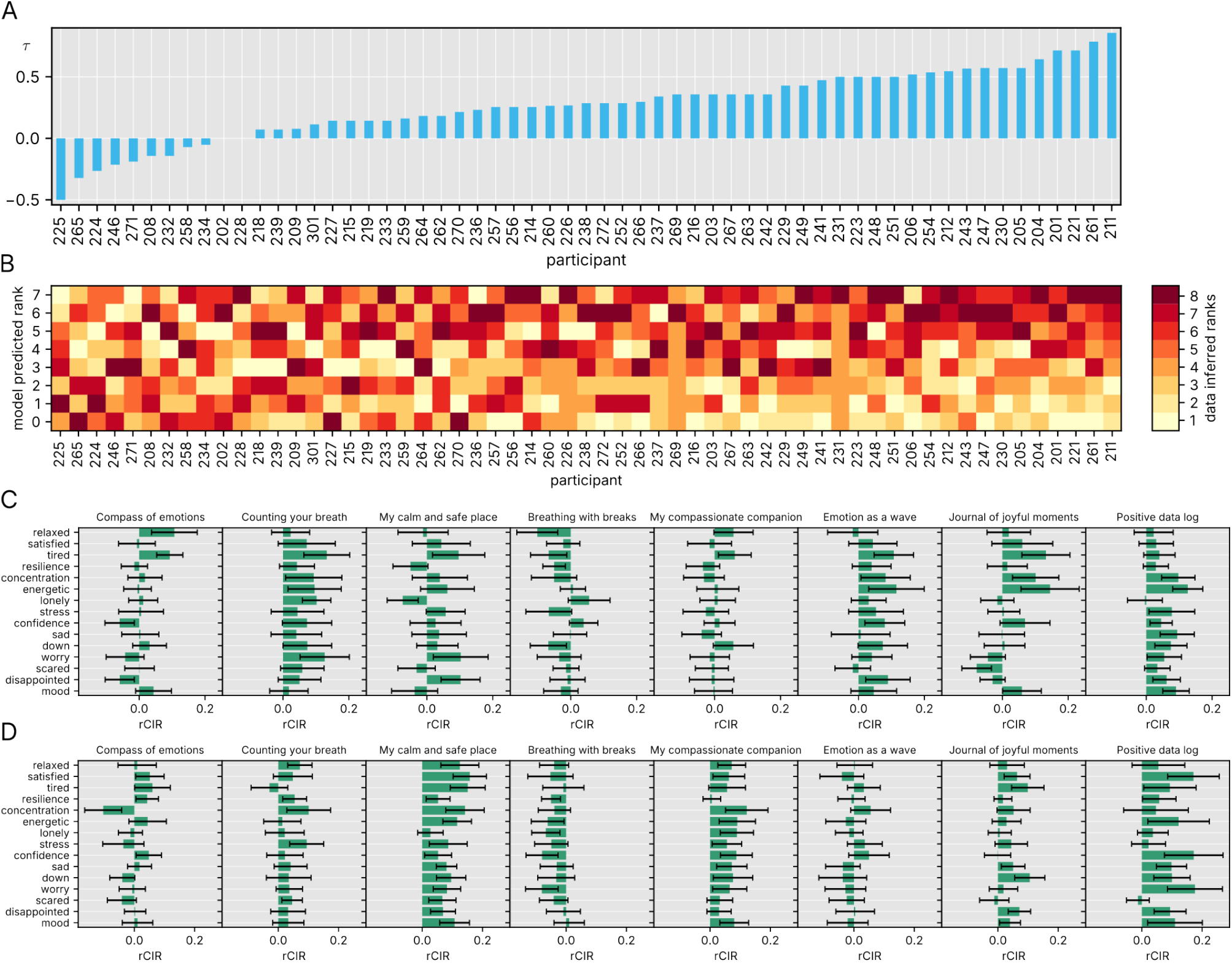
A: Rank correlation between predicted and empirical EMI effectiveness, given by Kendall’s *τ* for each participants of sample 3. Participants are shown in the order of correlation. Predicted effectiveness was measured by rCIR_1_, while empirical effectiveness was assessed as the one-step difference in EMA trajectories after the occurrence of an EMI. B: Empirical EMI ranks sorted by predicted EMI ranks for each participant of sample 3, in the same order as A. C: rCIR_7_ for each EMI in sample 2. Mean and standard error are displayed. D: Same for sample 3.

While we cannot easily validate these forecasts, the one-step ahead predictions of EMI effects and ground truth proximal effects are positively correlated for most participants (70% in sample 2 and 81% in sample 3, Fig. 4A and Appx. Fig. B6A), indicating our ability to personalize predictions across all 8 interventions by identifying both the most effective option and the order for each individual (Fig. 4B and Appx. Fig. B6B).

### Forecasting Implications For Real-World Study Designs

Several findings also carry important implications for real-world study design. Nonlinear models such as the PLRNN retained high predictive accuracy even in the presence of missing data, underscoring their practical applicability (Appx. Fig. B4B). Notably, data completeness—defined as a high proportion of non-missing responses—proved more critical for PLRNN performance than the overall length of the time series (Appx. Fig. B4A,B). This relationship varied by model type; for example, the VAR(1) showed a stronger dependence on data length.

Forecasting performance also systematically varied by EMA item type, with psychological variables generally exhibiting higher predictive accuracy than physical ones. For instance, the MAE for tired - which arguably reflects a more physical or somatic state - was approximately twice that of resilience (Appx. Fig. B3C). The rank ordering of prediction errors was nearly identical for the PLRNN and Kalman filter models, with only two items differing in rank (see Fig. B3A-C).

Moreover, these patterns were once more highly stable across samples, with Spearman rank correlations of *ρ* = 0.925 (*p* < .001).

Consistent with expectations, non-stationary items negatively impacted forecasting (*t*(1483) = −2.62, *p* = .004 across both samples), highlighting the importance of adaptive methods, though non-stationarity remained relatively minor over our 40-day studies.

Finally, incorporating additional activity-related external inputs improved forecasting performance compared to using only EMI as inputs—an effect observed specifically for the PLRNN (*t*(36) = 4.02, *p* < .001 across both samples), but not significantly for other network models. These findings suggest that combining a flexible, nonlinear forecasting model with relevant contextual information can further enhance predictive accuracy.

## 3 Discussion

This study demonstrates, for the first time, that nonlinear dynamical models markedly outperform conventional linear network approaches in predicting momentary psychological states from EMA data, and that their ability to incorporate EMI as external inputs may establish a robust methodological platform for next generation adaptive digital mental health interventions. By leveraging a series of intensive longitudinal datasets collected under ecologically valid conditions, we demonstrate that nonlinear SSMs—particularly the PLRNN—outperform traditional linear approaches in predictive accuracy and interpretability.

A key insight emerging from our analysis is that psychological processes, when modeled as interacting elements within a dynamic network, yield both more accurate forecasts and more meaningful representations of underlying mental states. This finding supports a growing consensus that mental health cannot be adequately captured through static or linear models, and instead reflects complex, nonlinear interactions evolving over time [16, 44–46]. Importantly, these interactions are not only quantifiable but also exploitable: by embedding psychological states in a generative latent space, the PLRNN enables precise perturbation analyses that can identify high-impact intervention targets, laying the basis for model-based predictive control [25, 47].

Most existing EMA studies rely on linear models such as VAR(1) or hierarchical regressions, which, while accessible and interpretable, cannot capture nonlinear transitions or latent influences. In contrast, the PLRNN not only accommodates such complexities but does so with sufficient efficiency to allow for on-device deployment and real-time feedback—essential features for scalable mental health technologies. Our finding that prediction performance remained robust even under conditions of partial missingness further underscores the PLRNN’s utility in real-world settings, where data sparsity is common [48, 49].

Beyond prediction, the nonlinear models revealed interpretable psychological networks whose structure remained consistent across participants and datasets. These networks exhibited face-valid patterns—for instance, stress and relaxation showed mutual inhibition—while linear models such as VAR produced counterintuitive links. The PLRNN’s capacity to map such networks stems from its piecewise-linear structure, which allows for the reconstruction of locally linear approximations of psychological dynamics that are both mathematically tractable and behaviorally interpretable, presenting a methodological advancement in psychological network anaylsis [50].

Crucially, these network representations served to inform potential interventions. By systematically perturbing individual network nodes, we identified states such as “sad” and “down” as high-leverage points whose targeted modulation—e.g., through EMI components—elicited widespread effects across the system. The strength of these effects scaled with network centrality. Indeed, forecasts of real EMI effects across a 24-hour window aligned with these insights, showing that most EMIs had either positive or neutral predicted outcomes, with high consistency across independent datasets.

We also identified key determinants of prediction quality. Forecasting accuracy varied by item type, with psychological variables (e.g., mood, anxiety) showing higher predictability than physical ones (e.g., tiredness), likely due to stronger embedding within the internal psychological network. Moreover, data completeness, rather than time series length, proved more important for prediction—particularly in nonlinear models—highlighting a clear design recommendation for future studies: prioritize sampling consistency over duration.

Taken together, the findings of improved predictive accuracy and interpretable insights into predictive mechanisms pave the way for personalized feedback and intervention designs based on nonlinear SSMs. However, while this work represents a significant advance, future studies should extend these models to incorporate multimodal inputs (e.g., passive sensing, geospatial data, clinical metadata, or biophysiological data) [51–53], explore adaptive model updating in long-term deployments [48], and refine the integration of control theoretic principles for intervention optimization (e.g., [25]), perhaps employing reinforcement learning techniques [54]. Nevertheless, despite the constraints of limited data, our findings demonstrate that nonlinear SSMs offer a powerful approach for accurately forecasting psychological states and intervention effects, laying the groundwork for real-time model-predictive control of mental-health interventions.

## 4 Methods

### Data and study design

We analyzed data from three micro-randomized trials (samples 1–3; *N* ≈ 60 each) conducted in the living lab AI4U [36]. Each trial comprised a ten-day training phase—during which eight EMI exercises (see Appx. A.2) were administered twice daily alongside eight random EMA prompts per day — followed by a thirty-day assessment phase with six EMA prompts daily and interspersed EMI, yielding up to 260 EMA observations and 200 EMI per participant (actual sampling varied with voluntary extra EMA and EMI requests). The EMA protocol included 15 primary well-being items (1–7 Likert; reverse-scored as needed, see Table A1) and six auxiliary covariates; EMA and binary EMI streams were temporally aligned, smoothed using a Gaussian kernel (8h window, *σ* = 1.5h) to accommodate irregular sampling [55, 56], and mean-centered using only training-set statistics to prevent information leakage. See Appx. A.1 and A.2.1 for elaborate details on study protocol, design, and data preprocessing.

### Forecasting and model evaluation protocol

We adopted an evaluation framework mirroring a real-time pipeline, in which forecasting models are retrained each night on all data up to the first EMA of the day and used to predict the subsequent seven observations - amounting to one day - at the time of EMI selection [36]. Eligible test days were those occurring on or after day 11 with at least three completed EMAs including the first of the day. Data prior to the first EMA defined the training set, and that day’s observations comprised the test set. Models whose fitting procedures involved non-deterministic optimization were trained ten times with random initializations, and forecasts were based on the ensemble mean. Predictive accuracy was assessed by MAE on absolute scores (see Appx. A.4).

### Modeling

Hyperparameters for all forecasting models were optimized by grid search on sample 1 to minimize MAE, with candidate ranges chosen to keep model complexity commensurate with each subject’s maximum time-series length. These fixed settings were then applied without further tuning to samples 2 and 3 for all comparative analyses.

We evaluated a hierarchy of models spanning from static baselines (last-observation carry-forward, global intercept, and linear regression with external inputs) through linear dynamical frameworks (VAR(1) and Kalman filter) to nonlinear state-space approaches (PLRNN trained via generalized teacher forcing [40]) and an autoregressive Transformer. Full details of each model’s implementation, training protocol, and hyperparameter ranges are provided in Appx. A.5, A.6.

### Network mechanisms

To characterize how psychological states influence one another, we derived effective connectivity matrices for each model by computing the Jacobian of the expected observation at time *t* with respect to the observation at *t* − 1. In the VAR(1) model this is simply the coefficient matrix *A* [14], whereas for the Kalman filter we compute *∂*𝔼[*x*_*t*_]*/∂x*_*t*−1_ = *B A B*^+^ (with *B*^+^ being the Moore–Penrose pseudoinverse). For the PLRNN, ReLU activations partition the latent space into linear regions {Ω_*j*_}, each with local Jacobian 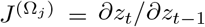; we then project these via 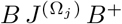 along each subject’s trajectory to obtain an average over time-varying connectivity.

To assess the impact of targeted interventions, we computed seven-step cumulative impulse responses (CIRs) for unit perturbations to each EMA node, defined as 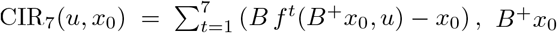 being the projection of the initial observation onto the latent space, and averaged across a set of representative initial conditions, explicitly including all standard basis vectors and the null vector. We then obtained the relative CIR (rCIR) by subtracting the autonomous response (CIR_7_(0, *x*_0_)) [57]. The sum of rCIRs across all variables (“total rCIR”) quantifies a node’s systemic influence, which we correlated with its out-degree centrality to identify high-impact targets for personalized interventions. See Appx. A.8 for more details.

### Determinants of forecasting performance

We first assessed item-level predictability by correlating each EMA item’s MAE and directional accuracy with its mean and stationarity using Spearman’s rank correlation. To characterize performance under limited data, we identified participants with at least ten eligible test days after time step 80 (17 in sample 2, 19 in sample 3), selected ten maximally spaced test days per subject, and trained models on pre-day windows of 30, 40, 50, 60, 70, and 80 time steps. We then simulated compliance rates from 80% down to 20% by randomly masking observations, repeating each scenario five times and averaging the outcomes. Finally, we evaluated the impact of four additional EMA covariates (sleep quality, quality of life, physical activity, activity pleasantness, see Appx. A.7) and two social-interaction items—normalized to [–1, 1] and appended to the input vector—by comparing model forecasts on the same low-data subsets with and without these extra inputs.

## Data Availability

All data produced in the present study are available upon reasonable request to the authors.

## Appendix A Supplementary Methods

### A.1 Sampling scheme

We analyzed three datasets from the AI4U living laboratory ([36]; www.ai4u-training.de). In each study iteration, 60 participants received various ecological momentary intervention (EMI) exercises on a smartphone and were prompted to complete ecological momentary assessment (EMA) surveys several times per day. After an initial screening, participants entered a 10-day training phase during which each of the eight EMI exercises was presented twice to familiarize them with the tasks. During this phase, eight EMA surveys were collected daily at random time points within a user-defined interval. Following the training phase, participants underwent 30 days of less intensive EMA sampling (six surveys per day). After each EMA, participants were asked if they wished to receive an EMI—termed “interactive tasks.” Additionally, one EMI was delivered daily at a user-defined time to ensure regular memory refreshing; these are termed “consolidation tasks.” At each delivery, the EMI was selected either randomly or via a selection algorithm (each with a 50% chance) from the set of eight possible interventions, with an option for a null intervention in which case no exercise was selected (see also [36]). Consequently, the expected maximum number of EMA timepoints per subject was 260 (detailed statistics on data completion are reported in Table B4). However, participants could voluntarily record additional EMAs, and in the second and third iteration, they could also request extra EMIs, leading to a variable number of EMAs and EMIs per day in practice. This study design is referred to as “micro-randomized trial” (sample). Consequently, we will refer to the study iterations as sample 1, sample 2, and sample 3 throughout the paper.

### A.2 EMA and EMI

During EMA assessment, a total of 17 items capturing constructs related to mental well-being were prompted. Of these, 2 were conditional, and only presented if an earlier item indicated the presence of negative emotions. Thus, the remaining 15 well-being items were selected for analysis. An additional 6 items measured external contexts and were used as covariates in a sub-analysis (see Section A.7). All 15 analyzed well-being items employed a 1–7 Likert scale, and negatively coded items (e.g., “I feel sad”) were reverse-scored so that higher values consistently signified better mental well-being (see Table A1 for the full item list). The EMI were selected from a set of 8 options focused on resilience, emotion regulation, and stress reduction. The set of EMI used in this study comprised: Compass of emotions, Counting your breath, My calm and safe place, Breathing with breaks, My compassionate companion, Emotion as a wave, Journal of joyful moments, Positive data log [58].

#### A.2.1 Data preprocessing

For every subject *s*, the EMA time series were encoded in a matrix *X*_*s*_ ∈ ℝ^*T ×n*^, where *T* is the number of recorded time steps and *n* is the number of EMA items. In *X*_*s*_, the *i*^th^ column represents the time course of EMA item *i*, and the *t*^th^ row contains all EMA responses at time point *t*. For brevity, we omit the subscript *s* in the following discussion.

Similarly, EMI time series were encoded in a binary matrix *U* ∈ {0, 1} ^*T*×*m*^, where *m* (here 8) is the number of admissible EMI. An entry of 1 in row *t* and column *i* indicates that intervention *i* was delivered at time *t*. Because interactive tasks were delivered immediately after an EMA, the rows of *U* were aligned with those of *X*. Consolidation tasks, delivered independently of EMA, were assigned to the preceding EMA time point to maintain temporal alignment.

To address the challenges posed by irregularly spaced EMA assessments, we smoothed the ordinal EMA data before model estimation. Accordingly, at each time point *t*, each EMA item was replaced by a weighted sum of all values recorded within a 4-hour window centered on *t*. The weights were determined using a Gaussian kernel with a standard deviation of 1.5 hours, so that measurements closer to *t* have a higher influence. Because EMA prompts occurred at randomized intervals, the resulting irregular spacing can complicate modeling [55]; kernel smoothing effectively mitigates these issues while aligning the data with model assumptions [56].

**Table A1.** EMA items included in this study and their properties. All were answered on a 1-7 Likert scale. Items formulated in a way that implied that high values indicated low mental well-being were reversed prior to model training and analysis.

|  | Code | Item | Reversed | Target construct |
| --- | --- | --- | --- | --- |
| 1 | ‘mood’ | I feel good. | no | positive affect |
| 2 | ‘disappointed’ | I am disappointed in myself. | yes | self-esteem |
| 3 | ‘scared’ | I feel anxious. | yes | negative affect |
| 4 | ‘worry’ | I am pensive. | yes | worrying |
| 5 | ‘down’ | I feel down. | yes | negative affect |
| 6 | ‘sad’ | I feel sad. | yes | negative affect |
| 7 | ‘confidence’ | I feel confident. | no | self-esteem |
| 8 | ‘stress’ | I feel stressed. | yes | stress |
| 9 | ‘lonely’ | I feel lonely. | yes | social isolation |
| 10 | ‘energetic’ | I am full of energy. | no | activity level |
| 11 | ‘concentration’ | I have difficulty concentrating. | yes | stress |
| 12 | ‘resilience’ | I can handle all the difficulties I may encounter. | no | resilience |
| 13 | ‘tired’ | I am tired. | yes | activity level |
| 14 | ‘satisfied’ | I am satisfied. | no | positive affect |
| 15 | ‘relaxed’ | I am relaxed. | no | positive affect |

After smoothing, each EMA item was mean centered. Importantly, no information from the evaluation/test set was used during this process. The evaluation/test data were left unsmoothed and uncentered, and only the mean from the training set was added back to the model predictions to restore them to the original data domain, ensuring a strict separation between training and evaluation sets.

### A.3 Forecasting and model evaluation protocol

For inclusion in the evaluation/test sets, days had to meet the following criteria:

- The day was the 11th day or later (i.e., following the 10-day training phase, ensuring a minimum amount of training data as specified in the AI4U study protocol).
- The day contained at least 3 non-missing EMA entries, providing enough samples for reliable prediction evaluation.
- The first EMA of the day was non-missing, ensuring a robust model initialization.

The number of eligible days was proportional to the number of non-missing EMA entries and thus varied greatly across subjects. Sufficiently robust data was obtained from 46 subjects in sample 1, 52 subjects in sample 2, and 53 subjects in sample 3 (see Table B4).

We evaluated the predicted EMA scores in each test set (referred to as “absolute score” throughout).

### A.4 Prediction accuracy metrics

Our main performance metric was the mean absolute error (MAE) which measures the distance in absolute value (L1) between predicted and true values. The MAE is more suitable than the more conventional mean squared error, since it avoids disproportionately penalizing larger deviations that may not carry proportionally greater meaning for non-Gaussian, bounded quantities like Likert scale values [59, 60].

To account for the varying number of test sets for every subject, as well as the varying number of non-missing EMA per set, we averaged the MAE first over the non-missing time steps and then over all evaluation/test set of a subject. They were then statistically compared on the between-subject level. MAE for subject *s* is given by

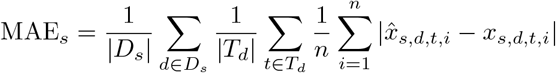

where *D*_*s*_ is the set of evaluation days for subject *s, T*_*d*_ is the set of non-missing EMA on day *d*, and 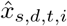 is the predicted value for EMA item *i* at time *t* on day *d* for subject *s*.

Secondly, we separately calculated the MAE for data points immediately proceeded by an EMI

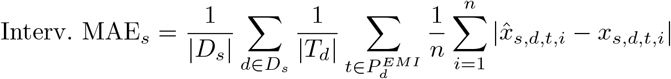

where *P*_*d*_ is the set of successive pairs (times *t* and *t* + 1) of EMA on day *d*, where both EMA are non-missing, and *u*_*t*_ ≠ 0, i.e. there was an intervention at time *t*.

### A.5 Modeling

We evaluated a series of forecasting models of increasing complexity, allowing us to systematically isolate and assess the contributions of various methodological components, focusing on dynamic network-type models. Our approach started with the simple, non-dynamic models, and then progressively incorporated additional features—such as external inputs, latent representations, and non-linear recurrence—to more accurately capture the underlying dynamics of the EMA time series. Ultimately, the list of tested models comprised:

- static models, including
  - the last time step,
  - the global intercept, and
  - a linear regression model including an intercept and external inputs,
- dynamic models, including
  - vector autoregressive models of order 1 (VAR(1)),
  - linear state space models (SSMs) with latent vector autoregressive component of order 1 (also known as a Kalman filter),
  - non-linear SSMs (with a piecewise linear recurrent neural network (PLRNN) as latent model), and
  - autoregressive Transformer models.

Whereas VAR models are commonly used in EMA analysis [14], the PLRNN trained via generalized teacher forcing (GTF) is state-of-the-art (SOTA) in the reconstruction of benchmark and real-world dynamical systems [40, 60, 61], whereas Transformers are currently among the most powerful AI algorithms for modeling complex sequential data [43, 62]. We included the static models as naive yet powerful baselines that have been shown to occasionally outperform even recent SOTA time series models, probably due to overfitting [63].

In the following, we describe each model in detail, emphasizing the progressive increase in complexity to facilitate direct comparability.

#### Last time step

In n-step ahead prediction, we use the value of the last time step as a predictor, modeling

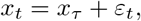

for *t* = *τ* + 1, …, *τ* + 7, where *τ* is the last time point of the training data and *ε*_*t*_ represents the error made at time *t*.

#### Global intercept

This model predicts the next value as the average over all previous observations:

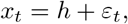

where this average is given by 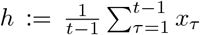. Both the last time step and the global intercept models ignore external inputs and predict constant time series; consequently, they cannot forecast temporal changes within a day.

#### Linear regression model

By incorporating external inputs into the global intercept model, we obtain a multivariate linear regression model of the form

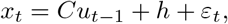

where *h* ∈ ℝ^*n*^ is a bias (offset) v ector, *C* ∈ ℝ^*n×m*^ i s t he r egression c oefficient mat rix map ping the external inputs *u*_*t*−1_ to the EMA space, and *ε*_*t*_ is a normally distributed error. Although this model still lacks a recurrent (autoregressive) component, it allows dynamic predictions due to time-varying external inputs.

#### Linear autoregressive model

Including the EMA variables from preceding time steps as linear predictors in addition to the external inputs leads to the VAR(1) model of the form

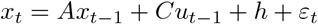

with *A* ∈ ℝ^*n×n*^. EMA variables *x*_*t*_ are now modeled as a recurrent variable that changes over time in an autoregressive fashion. Matrix *A* encodes the linear influence of *x* at time *t ™* 1 on itself at time *t*.

The VAR(1) model (a linear dynamical system) is currently the most widespread model in the analysis of EMA time series data [14]. The model has *n*^2^ + *nm* + *n* free parameters (Θ = {*A, C, h*}) and no hyperparameters. Being essentially a linear regression model, the likelihood has a closed-form solution, making it straightforward to infer. In this study, we used ridge regression to ensure that the absolute maximum eigenvalue |*λ*_max_(*A*) | remained less than 1, thereby ensuring stability [64].

#### Linear SSM

An extension of the VAR(1) model involves representing the autoregressive process within a latent space, yielding a state-space model (SSM). This latent vector autoregressive model—commonly referred to as the Kalman filter [65]—is given by

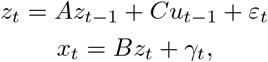

where *ε*_*t*_ ~ 𝒩 (0, Σ), *γ*_*t*_ ~ 𝒩 (0, Γ), and *z*_0_ ~ 𝒩 (*µ*_0_, Σ). Here, the recurrent variable is *z*, which represents an unobserved latent process from which observations *x* are derived. As a result, once we condition on the latent state *z*, the EMA variables are treated as conditionally independent. This additional flexibility enables the SSM to capture latent states that are not directly observed but critical for the system’s dynamics.

The only hyperparameter is the size of the latent state, *l*. The model has *l*^2^ + *nl* + *ml* free parameters (Θ = {*A, B, C*}), which in our case is less than the VAR(1) model if *l* ≤ 10. The Kalman filter was inferred using a version of the Expectation Maximization (EM) algorithm [66–68] that guaranteed non-divergence in the latent process (by |*λ*_max_(*A*) | *<* 1). Since the EM algorithm can lead to exponentially diverging parameter estimates when initialization is suboptimal or when data are scarce, we mitigated this risk by performing up to 10 different random initializations for each model, thereby increasing the chance of convergence.

#### Nonlinear SSM

Replacing the linear recurrence of the Kalman filter with a non-linear function such as the rectified linear unit (ReLU), tanh, or sigmoid results in a recurrent neural network (RNN) SSM. This opens the door to a class of highly powerful models that, in principle, are capable of emulating the dynamics of any real-world system [69]. In this study, we utilize an RNN-based SSM that has proven highly effective i n l earning a nd reconstructing nonlinear dynamical systems across a range of benchmarks and data modalities, due to its superior SOTA training algorithm [39–41, 61, 70]. We employed the two-layered version of [40] called ‘shallow PLRNN’. The model is given by

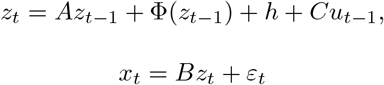

with nonlinearity

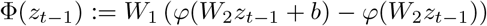

where *z*_*t*_ ∈ ℝ^*l*^ is the latent state, *A* ∈ ℝ^*l×l*^ is a diagonal matrix, *W*_1_ ∈ ℝ^*l×k*^ and *h* ∈ ℝ^*l*^ are weight matrix and bias of layer 1, respectively, and *W*_2_ ∈ ℝ^*k×l*^ and *b* ∈ ℝ^*k*^ are weight matrix and bias of layer 2. The rectified linear unit activation functions

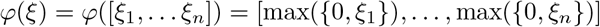

partitions the state space into linear subregions. Nevertheless, the PLRNN is a universal function approximator [69, 71, 72]. Matrix *C* ∈ ℝ^*l×m*^ maps the external inputs into the latent space. Matrix *B* ∈ ℝ^*n×l*^ the latent states onto the predicted EMA values (analogous to the Kalman filter).

Free model parameters are Θ = {*A, W*_1_, *W*_2_, *h, b, B, C*}, altogether 2*lk* + 2*l* + *k* + *ln* + *lm* in number. The size of the latent states, *l*, the size of the second layer *k*, learning rate, teacher forcing strength *α*, batch size, and sequence length were hyperparameters optimized via a gridsearch on the data from sample 1.

The PLRNN was trained with backpropagation through time on sequences of length *τ* drawn from a subject’s time series, where *τ* was a hyperparameter. During each iteration, mean squared error loss was calculated for batches of size *k* (also a hyperparameter). To prevent exploding gradients, we used GTF [40]: during training, and after evaluating the loss, the latent states are replaced by a weighted sum of *z*_*t*_ and a back-projection of the data into the latent space, the so-called forcing signal,

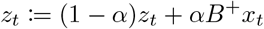

where 0 ≤ *α <* 1 and *B*^+^ is the Moore-Penrose pseudoinverse of *B*. In case that *x*_*t*_ was missing, *z*_*t*_ was left unchanged.

**Table A2.** Optimized hyperparameters and resulting total number of free parameters of all models. Only applicable hyperparameters are shown.

| Hyperparameter | Linear Regr. | VAR(1) | Kalman filter | PLRNN | Transformer |
| --- | --- | --- | --- | --- | --- |
| latent dim |  |  | 7 | 6 | 4 |
| hidden/FC layer dim |  |  |  | 15 | 5 |
| # encoder layers |  |  |  |  | 1 |
| # decoder layers |  |  |  |  | 1 |
| # heads |  |  |  |  | 1 |
| initial learning rate | | | | $10^{-3}$ | $10^{-3}$ |
| learning rate annealing |  |  |  | on | off |
| batch size |  |  |  | 16 | 16 |
| sequence length |  |  |  | 7 | 32 |
| TF strength $\alpha$ | | | | 0.125 | |
| # free parameters | 135 | 360 | 210 | 300 | 645 |

#### Autoregressive Transformer

Even though the Transformer neural network has only an indirect representation of time via positional encoding, it can be used in an autoregressive fashion to predict future time steps of a time series given its past and covariates (e.g. [73, 74]). It consists of an encoder and a decoder block with *K*_enc_ and *K*_dec_ layer, respectively, identical to those used in [42]. Each has *h* attention heads. The encoder outputs a latent state of size *M*, which is fed into the decoder, along with the last *τ* time steps of the input time series, which are then predicted 1 step into the future, using masked attention [42]. It then outputs the next predicted time step.

In contrast to the methods above, the Transformer is not an extension of the previously introduced models, but has an entirely different architecture. It also lacks the Markov property and always needs the complete past time series to perform predictions.

### A.6 Hyperparameter optimization

Some of the models we employed include hyperparameters that require careful tuning. To avoid the double-dipping problem, hyperparameter optimization with MAE as target was conducted using the data from sample 1, while all subsequent model analyses were conducted using the data from sample 2 and sample 3. Hyper-parameters were optimized via grid search, systematically testing all combinations of several candidate values for each hyperparameter. Optimal hyperparameter settings (see table A2) resulted in different numbers of free parameters. Due to the encoder/decoder structure, and every layer containing an additional fully connected layer, a transformer cannot get much smaller than used here.

### A.7 Determinants of forecasting ability

#### Item-level forecasting accuracy

Apart from evaluating the overall forecasting ability averaged over EMA, we analyzed the predictability of individual EMA items. We tested for contingencies of item accuracy metrics with item mean, item variance, and stationarity using Spearman rank correlations.

#### Low data limit

A central and empirically relevant question that arises is how prediction quality increases with available data. To address this, we selected the participants that - after time step 80 - had at least 10 eligible test days. We identified 20 participants in sample 2 and 17 participants in sample 3. To equalize the amount of test data, we selected exactly 10 maximally spread-out (to remove auto-correlations) test days for each subject. We then trained models on the 30, 40, 50, 60, 70, and 80 steps preceding each test day, respectively, to simulate different amounts of training data.

**Table A3.**
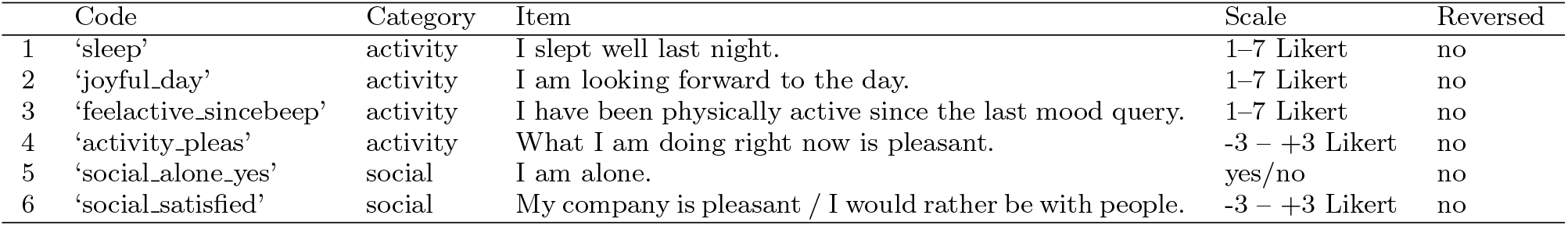
EMA items used as additional external inputs in sub-analysis A.7. Items 2 and 3 were only queried if item 1 was answered “yes”, i.e. on the first EMA of the day.

All of these subjects had valid data point ratios of *>* 60%. We simulated lower compliance rates by randomly setting time points to NaN, creating data sets with valid ratios of at most 80%, 70%, 60%, 50%, 40%, 30%, and 20%. We then repeated the analysis five times and averaged the results.

#### Additional covariates

Finally, we analyzed whether including additional external covariates, other than EMIs, would improve prediction. These comprised some of the EMA items not used as dependent variables, as listed in Table A3. We normalized the Likert items to the [−1, 1] interval, and encoded binary variables as 0/1, to align their magnitudes with those of the (binarily encoded) EMI. These inputs were appended to the input vectors *u*_*t*_. To evaluate the impact of the additional covariates, we trained the models with and without the 4 additional EMA items encoding sleep quality, quality of life, physical activity, and pleasantness of the current activity, and with and without the 2 additional EMA items encoding social interactions. The same subjects and prediction days as in A.7 were used for training.

### A.8 Network mechanisms

#### Network connectivity

One major advantage of the VAR(1) model in EMA analysis is its straightforward interpretation as an ideographic network model, where psychological states directly influence each other over time. In a VAR(1) model, the derivative of *x*_*t*_ with respect to *x*_*t*−1_ is exactly 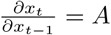, meaning each element *A*_*ji*_ clearly represents the linear influence of variable *i* on variable *j* [14]. In contrast, in a Kalman filter the state transition matrix *A* governs the dynamics of the latent state *z*_*t*_, making it less straightforward to interpret. However, by obtaining the derivative of *x*_t_ with respect to *x* as 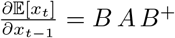, with *B*^+^ being the Moore-Penrose pseudoinverse of *B*, we can translate the latent state connectivity into an effective connectivity among the observed variables. In the PLRNN, the ReLU activation partitions the latent space into a set of subregions Ω_1_, …, Ω_*n*_. Inside each subregion Ω_*j*_, the dynamics are linear, so for any *z*_*t*−1_ ∈ Ω_*j*_ the evolution of the latent state is governed by the local Jacobian 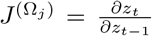 if *z*_*t*−1_ ∈ Ω_*j*_. Hence, the derivative of observed variable *x*_*t*_ w.r.t. *x*_*t*−1_ is 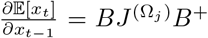, where Ω is determined by the location of *B*^+^*x* in the latent space. Thanks to the piecewise linearity of the model and its Markov property, the local Jacobian can be computed analytically. We used this idea to visualize the PLRNNs as ideographic networks. To obtain a realistic representation of the networks, we computed the Jacobians along the trajectory of EMA variables *X* for each participant (based on the model trained on the longest available train set with lowest test loss).

#### Node centrality

The centrality of a network node *i* was computed as its weighted absolute out-degree centrality. Let *V* be the set of nodes and *e*_*ij*_ the weight of the edge from node *i* to node *j*, then it is defined as

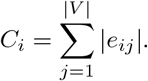

#### Cumulative impulse response

The cumulative impulse response (CIR) quantifies the overall effect that an external input *u* has on a system’s observations over time [57]. Each entry in the CIR vector corresponds to the effect on the respective observed variable. In a PLRNN, it is defined as

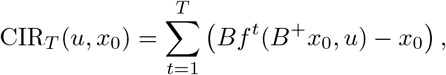

where *f*^*t*^ denotes the state of the system after *t* iterations, and *z*_0_ is the initial condition. Because the system is nonlinear, the response CIR_*T*_ (*u, z*_0_) can vary significantly for different initial states. To ensure that our measure of the system’s response is robust and not overly influenced by a particular initial condition, we average the CIR over many representative starting points. Specifically, we evaluated CIR_*T*_ (*u, e*_*j*_) for each standard basis vector *e*_*j*_ plus the null vector and then compute the mean. We set *T* = 7 to mirror the length of our test sets (24 hours). To separate the effects of *u* from the autonomous dynamics, we defined the relative CIR (rCIR) as CIR_7_(*u, z*_0_) − CIR_7_(0, *z*_0_), i.e. CIR minus the cumulative trajectory without external influence. This targeted perturbation allows us to isolate the influence of each variable, thereby shedding light on its unique role within the causal network, and potentially uncover therapeutic targets.

We evaluated *r*CIR_7_(*u, x*_0_) using external inputs *u* that were specifically designed to affect only one EMA variable at a time. This was achieved by determining a basis for the orthogonal complement of ker(*B*_*j*_ · *C*), where *B*_*j*_ was a single row of the observation matrix *B*, corresponding to the EMA variable in question. Note that the orthogonal complement is at most 1-dimensional; the 0-dimensional case did not occur. By choosing *u* as the normalized (length 1) basis vector, we made sure it was targeted at the specific variable with normed input strength.

## Appendix B Supplementary Results

### B.1 Data characteristics

For each sample, 60 participants were recruited. Only participants who completed the data collection phase were included, resulting in *n* = 57 subjects in sample 1, *n* = 56 in sample 2 and *n* = 59 in sample 3, respectively. Models were trained only on those participants who had at least one day that met the criteria for test set inclusion (*n* = 46 subjects in sample 1, *n* = 48 in sample 2 and *n* = 51 in sample 3, see section A.3). Table B4 provides details on the number and proportion of non-missing EMA entries, as well as the number of EMI instances. Across these key data characteristics, sample 1, 2 and 3 were nearly identical. Furthermore, the distributions of all EMA items did not significantly differ between the samples (all *p* > .9 in a Kolmogorov-Smirnov test). This supported the assumption that the optimal hyperparameters for models trained on sample 2 and 3 would also be the same.

**Table B4.** Essential data characteristics. Median and 25th/75th percentile are reported.

|  | sample 1 | sample 2 | sample 3 |
| --- | --- | --- | --- |
| # subjects | 57 | 56 | 59 |
| # subjects included | 46 | 48 | 51 |
| # non-missing EMA per subject | 115.5 (92.5, 175) | 125.5 (102.5, 152) | 127 (90, 160) |
| % non-missing EMA per subject | 48.2 (25.4, 68.9) | 49.6 (40.7, 59.9) | 49.6 (35.7, 63.5) |
| # EMI per subject | 61.5 (46.25, 88) | 65 (47, 93.25) | 61 (34, 87) |
| # of eligible days per subject | 6 (3, 17.5) | 7.5 (3, 14) | 7 (3, 17) |

**Figure B1.**
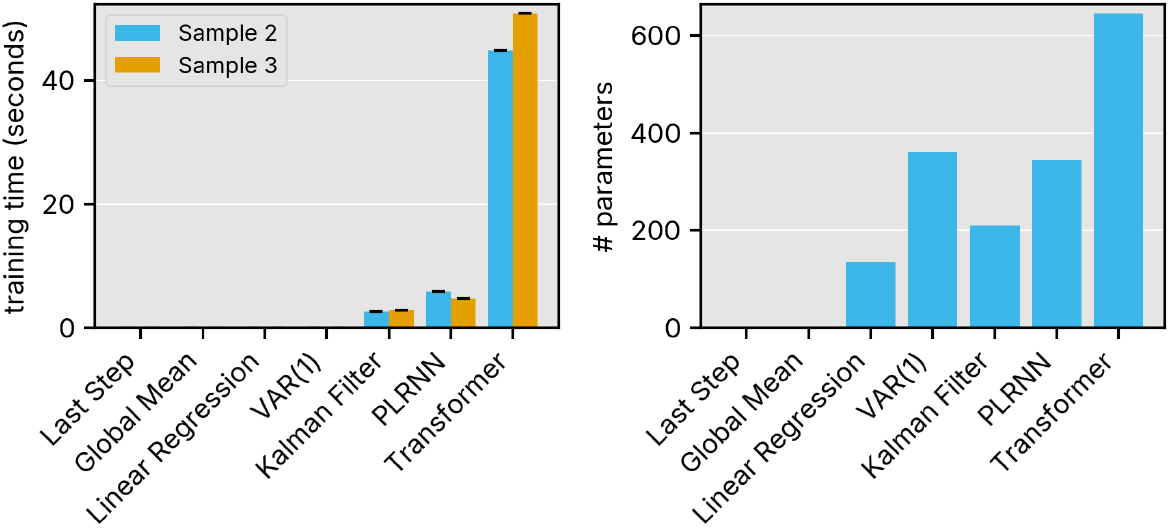
Training time (left) and number of parameters (right) for each model. Results show means across participants with bootstrapped confidence intervals.

Item stationarity was assessed using the augmented Dickey-Fuller test [75]. The null hypothesis that the time series were non-stationary was rejected for *p < α* = 0.05. For most subjects (82% in sample 1, 77% in sample 2, 80% in sample 3), at least 14 out of 15 EMA items were stationary.

### B.2 Additional analyses

#### Model training effort

In real-world scenarios, saving compute time and memory is crucial. Hence, we compared the models in terms of training time (for a single model) and number of parameters. Last step, global mean, linear regression, and VAR(1) models have closed-form optimal solutions, requiring only fractions of a second to train per model. As depicted in Fig. B1, Kalman filters took less than 5 seconds and PLRNNs less than 8 seconds to train on average. The up to 50 seconds needed to train a Transformer are a huge disadvantage for the deployment in a real-time application. Models were trained on a 32-core AMD Epyc 7452 server with 1 TB RAM (however, not more than 1 GB of RAM was occupied at a single time).

The higher parameter count in PLRNN and Kalman filter models reflects their latent structure with freely scalable latent dimensions, in contrast to VAR(1), which must learn dynamics directly in the 15-dimensional observation space. Transformers also had over 1.5 times more parameters compared to PLRNNs and over 3 times more than Kalman filters in the optimal hyperparameter configuration.

**Figure B2.**
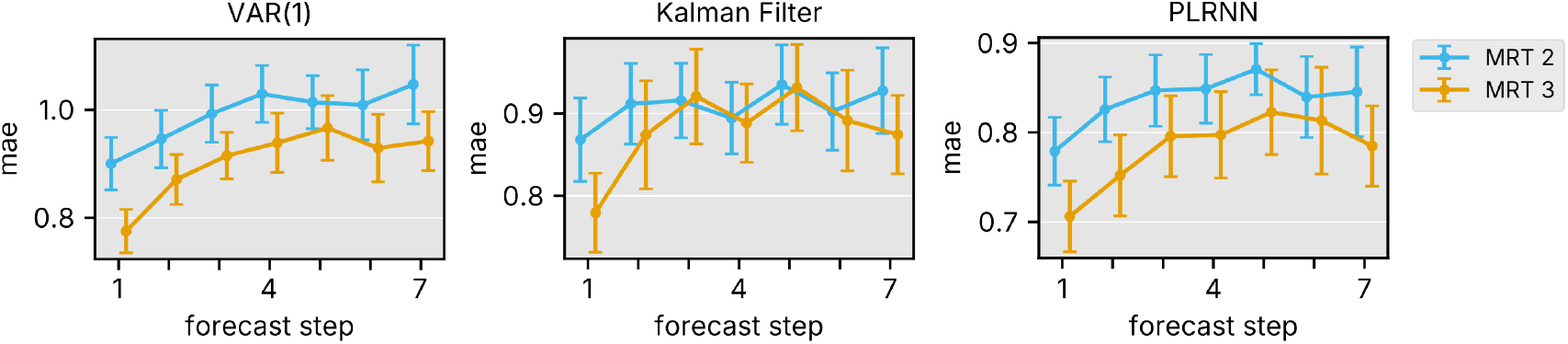
Stepwise mean absolute error (MAE) of predicted absolute EMA values for the VAR(1) model (left), Kalman filter (middle), and PLRNN (right). The x-axis indicates the prediction step relative to the start of the test set. Results show means across participants with bootstrapped confidence intervals.

**Figure B3.**
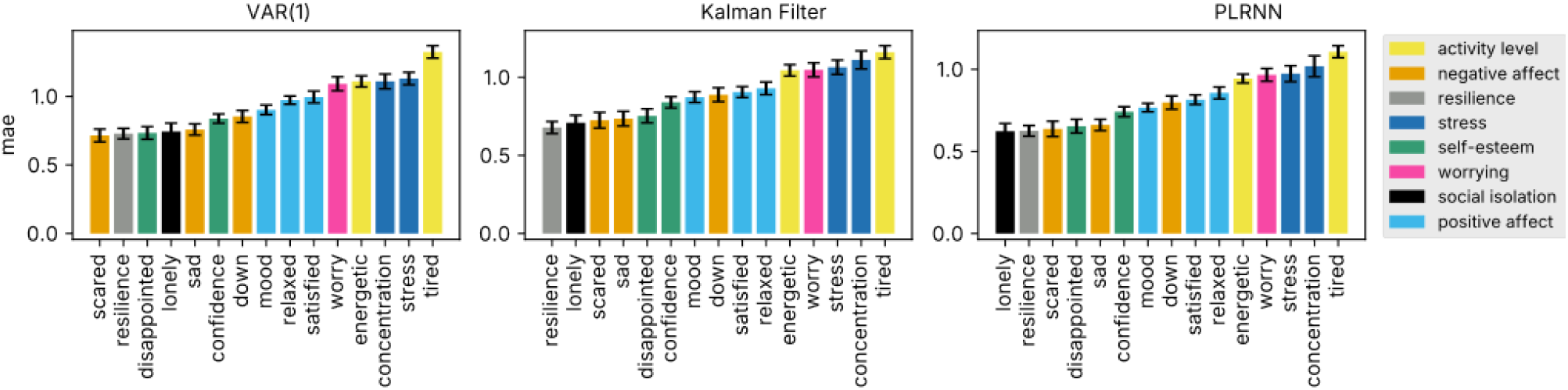
MAE for the absolute score of each EMA item for VAR(1), Kalman filter, and PLRNN. Mean over participants from both samples and standard errors are displayed. The colors indicate the psychological constructs the EMA items refer to.

#### Stepwise prediction accuracy

When breaking down the MAE by individual forecasting steps, it becomes evident that forecasting EMA values becomes progressively more difficult the further ahead the forecast reaches (see Fig. B2). This pattern aligns with expectations and provides a basic validation of the model’s temporal behavior.

#### Individual EMA items

EMA items differed substantially in their predictability as measured by MAE. For example, the item ‘tired*’ was about twice as hard to predict as the items ‘lonely*’ and ‘resilience*’. See Fig. B3 for details.

#### Low data limit

We analysed forecasting performance under sparse data conditions by systematically removing time steps from the beginning of the train set for selected participants. While the PLRNN reached its maximum predictive accuracy with as few as 30 steps, the Kalman filter required at least 50 steps. In contrast, the VAR(1) model showed a near-linear improvement with increasing time series length (see Fig. B4A). In a second analysis, we compared the empirical (full) time series length of all participants with prediction performance. The above observation was reflected here: the MAE tended to decrease linearly with longer train sets for the VAR(1) model (*p <* .001), but not for the Kalman filter (*p* = .864) nor the PLRNN (*p* = .108, Bonferroni corrected for 3 tests).

**Figure B4.**
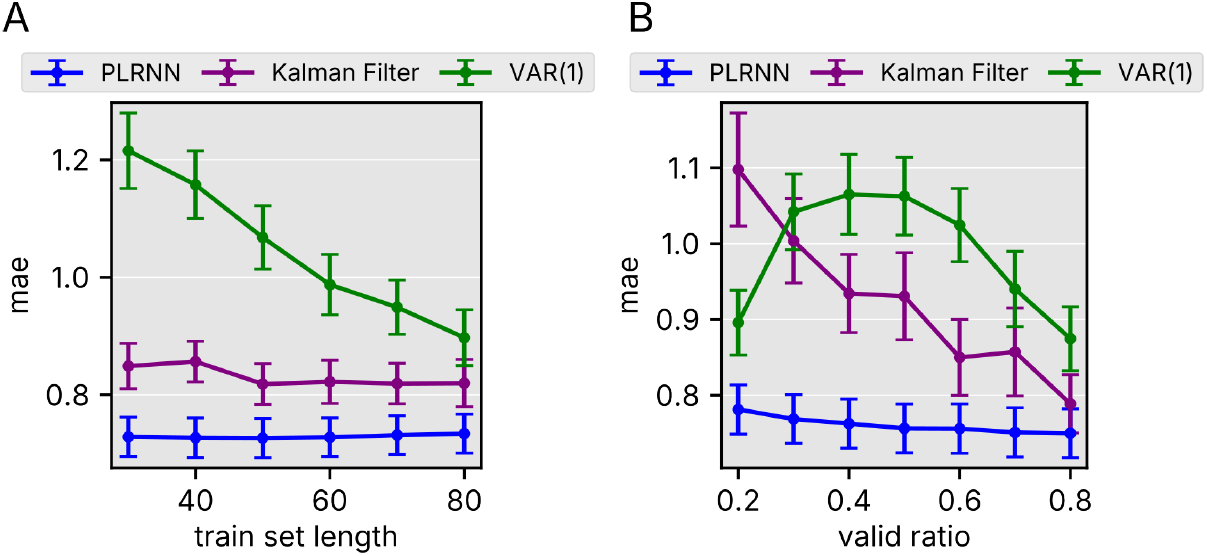
MAE of predicted absolute scores for different determinants of forecasting quality. A: MAE over train set length. Different lengths were produced by removing time steps from the beginning of the time series. Mean over participants and standard error are displayed. B: MAE over ratio of valid data in the train set. Valid ratios were produced by randomly setting steps from the time series to NaN. Mean over participants and standard error are displayed.

**Figure B5.**
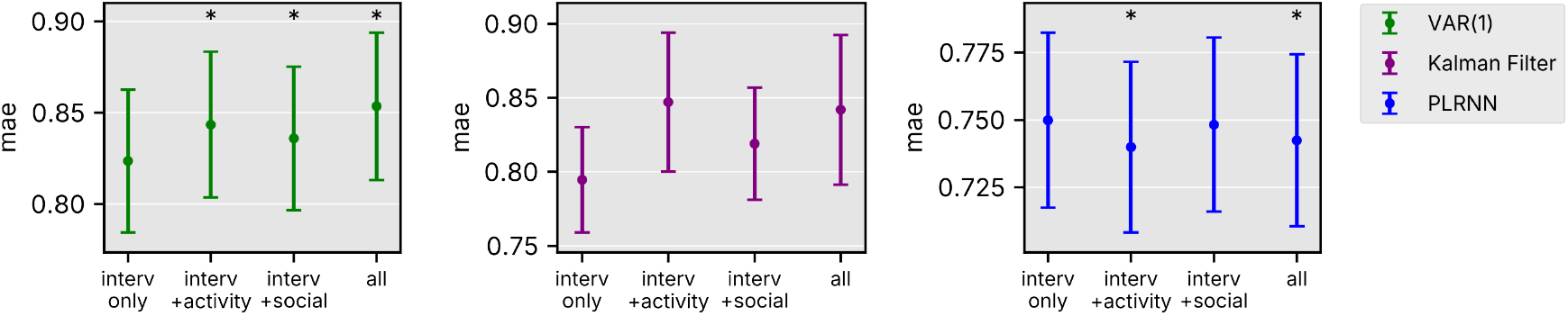
MAE for the VAR(1) model (left), Kalman filter (middle) and PLRNN (right), as a function of additional covariates (Interv only = interventions used as external inputs, no additional covariates; interv+activity = additional activity-related EMA included as inputs; interv+social = additional social-related EMA included as external inputs; all = all additional EMA included as external inputs). Asterisks indicate significant differences compared to the *interv only* condition, based on paired t-tests.

In a similar manner, we emulated specific levels of missingness by removing random time steps from the train set of selected participants. Both the PLRNN and the Kalman filter exhibited a reduced MAE for higher ratios of valid (non-missing) data. Notably, the PLRNN achieved low prediction errors with as little as 20% valid data. Interestingly, the VAR(1) model performed relatively well under high levels of missingness, exhibiting its highest error around 40% valid data, followed by a roughly linear decrease thereafter. Results are displayed in Fig. B4B. Consistent with these findings, prediction performance across all participants improved with increasing empirical valid data ratio for all three models (all *p <* .001, Bonferroni corrected for 3 tests).

#### Additional covariates

While the PLRNN benifited significantly from the inclusion of activity-related EMA items as external regressors but not social-related items, including both types also increased performance (*t*(36) = 2.83, *p* = .008).

#### Prediction of EMI ranks for sample 2

For completeness, we present the prediction of EMI ranks for sample 2, akin to Fig. 4A, B for sample 3 (main manuscript).

**Figure B6.**
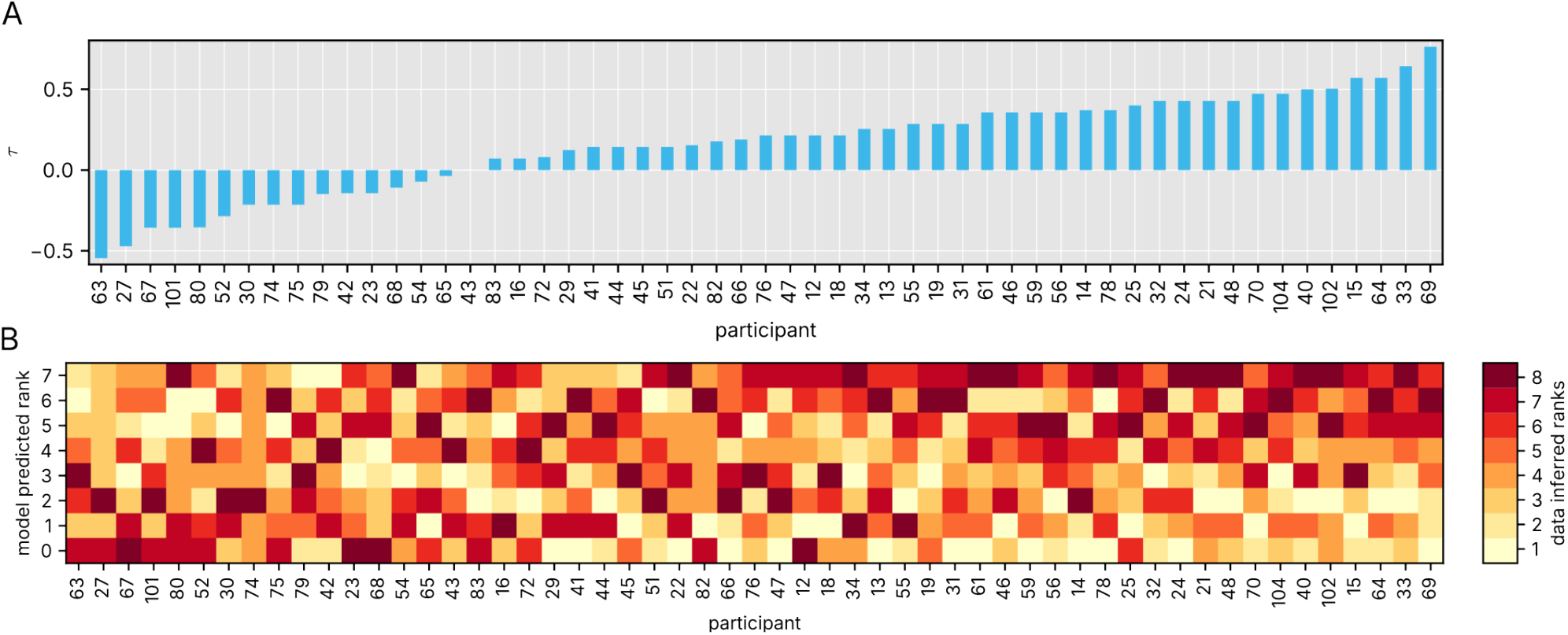
A: Rank correlation between predicted and empirical EMI effectiveness, given by Kendall’s *τ* for each participants of sample 2. Participants are shown in the order of correlation. Predicted effectiveness was measured by CIR_1_, while empirical effectiveness was assessed as the one-step difference in EMA trajectories after the occurrence of an EMI. B: Empirical EMI ranks sorted by predicted EMI ranks for each participant of sample 2, in the same order as A.

## References

[1] Donker, T. et al. Smartphones for Smarter Delivery of Mental Health Programs: A Systematic Review. Journal of Medical Internet Research 15, e247 (2013).

[2] Firth, J. et al. The efficacy of smartphone-based mental health interventions for depressive symptoms: a meta-analysis of randomized controlled trials. World Psychiatry 16, 287–298 (2017).

[3] Neben, T. et al. Make the Most of Waiting: Theory-Driven Design of a Pre-Psychotherapy Mobile Health Application. In AMCIS 2016 Proceedings (2016).

[4] Huang, S., Wang, Y., Li, G., Hall, B. J. & Nyman, T. J. Digital Mental Health Interventions for Alleviating Depression and Anxiety During Psychotherapy Waiting Lists: Systematic Review. JMIR Mental Health 11, e56650 (2024).

[5] Erbe, D., Eichert, H.-C., Riper, H. & Ebert, D. D. Blending Face-to-Face and Internet-Based Interventions for the Treatment of Mental Disorders in Adults: Systematic Review. Journal of Medical Internet Research 19, e306 (2017).

[6] Schick, A. et al. Effects of a Novel, Transdiagnostic, Hybrid Ecological Momentary Intervention for Improving Resilience in Youth (EMIcompass): Protocol for an Exploratory Randomized Controlled Trial. JMIR Research Protocols 10, e27462 (2021).

[7] Shiffman, S., Stone, A. A. & Hufford, M. R. Ecological Momentary Assessment. Annual Review of Clinical Psychology 4, 1–32 (2008).

[8] Nahum-Shani, I. et al. Just-in-Time Adaptive Interventions (JITAIs) in Mobile Health: Key Components and Design Principles for Ongoing Health Behavior Support. Annals of Behavioral Medicine 52, 446–462 (2018).

[9] Myin-Germeys, I. et al. Experience sampling methodology in mental health research: new insights and technical developments. World Psychiatry 17, 123–132 (2018).

[10] Schick, A. et al. Novel digital methods for gathering intensive time series data in mental health research: scoping review of a rapidly evolving field. Psychological Medicine 53, 55–65 (2023).

[11] Heron, K. E. & Smyth, J. M. Ecological momentary interventions: incorporating mobile technology into psychosocial and health behaviour treatments. British Journal of Health Psychology 15, 1–39 (2010).

[12] Myin-Germeys, I. et al. Experience sampling research in psychopathology: opening the black box of daily life. Psychological Medicine 39, 1533–1547 (2009).

[13] Borsboom, D. A network theory of mental disorders. World Psychiatry 16, 5–13 (2017).

[14] Bringmann, L. F. et al. A Network Approach to Psychopathology: New Insights into Clinical Longitudinal Data. PLOS ONE 8, e60188 (2013).

[15] Hamaker, E. L., Ceulemans, E., Grasman, R. & Tuerlinckx, F. Modeling affect dynamics: State of the art and future challenges. Emotion Review 7, 316–322 (2015).

[16] Durstewitz, D., Huys, Q. J. M. & Koppe, G. Psychiatric Illnesses as Disorders of Network Dynamics. Biological Psychiatry: Cognitive Neuroscience and Neuroimaging 6, 865–876 (2021).

[17] Vallacher, R. R., Van Geert, P. & Nowak, A. The Intrinsic Dynamics of Psychological Process. Current Directions in Psychological Science 24, 58–64 (2015).

[18] Hollenstein, T., Lichtwarck-Aschoff, A. & Potworowski, G. A Model of Socioemotional Flexibility at Three Time Scales. Emotion Review 5, 397–405 (2013).

[19] Marks-Tarlow, T. The Self as a Dynamical System. Nonlinear Dynamics, Psychology, and Life Sciences 3, 311–345 (1999).

[20] Scheffer, M. et al. A Dynamical Systems View of Psychiatric Disorders—Practical Implications: A Review. JAMA Psychiatry 81, 624 (2024).

[21] Meine, L. E. et al. Network analyses of ecological momentary emotion and avoidance assessments before and after cognitive behavioral therapy for anxiety disorders. Journal of Anxiety Disorders 106, 102914 (2024).

[22] Tseng, W.-L. et al. Network analysis of ecological momentary assessment identifies frustration as a central node in irritability. Journal of Child Psychology and Psychiatry 64, 1212–1221 (2023).

[23] Salvi, J. D., Rauch, S. L. & Baker, J. T. Behavior as Physiology: How Dynamical-Systems Theory Could Advance Psychiatry. American Journal of Psychiatry 178, 791–792 (2021).

[24] Gauld, C. & Depannemaecker, D. Dynamical systems in computational psychiatry: A toy-model to apprehend the dynamics of psychiatric symptoms. Frontiers in Psychology 14, 1099257 (2023).

[25] Fechtelpeter, J. et al. A control theoretic approach to evaluate and inform ecological momentary interventions. International Journal of Methods in Psychiatric Research 33, e70001 (2024).

[26] Myin-Germeys, I., Klippel, A., Steinhart, H. & Reininghaus, U. Ecological momentary interventions in psychiatry. Current Opinion in Psychiatry 29, 258–263 (2016).

[27] Schulte-Strathaus, J. C. C., Rauschenberg, C., Baumeister, H. & Reininghaus, U. Ecological Momentary Interventions in Public Mental Health Provision. In Digital Phenotyping and Mobile Sensing: New Developments in Psychoinformatics (eds Montag, C. & Baumeister, H.) Studies in Neuroscience, Psychology and Behavioral Economics, 427–439 (Springer International Publishing, Cham, 2023).

[28] Balaskas, A., Schueller, S. M., Cox, A. L. & Doherty, G. Ecological momentary interventions for mental health: A scoping review. PLOS ONE 16, e0248152 (2021).

[29] Pooseh, S., Kalisch, R., Köber, G., Binder, H. & Timmer, J. Intraindividual time-varying dynamic network of affects: linear autoregressive mixed-effects models for ecological momentary assessment. Frontiers in Psychiatry 15, 1213863 (2024).

[30] Terhorst, L. et al. Hierarchical Linear Modeling for Analysis of Ecological Momentary Assessment Data in Physical Medicine and Rehabilitation Research. American Journal of Physical Medicine & Rehabilitation 96, 596–599 (2017).

[31] Kim, J., Marcusson-Clavertz, D., Togo, F. & Park, H. A Practical Guide to Analyzing Time-Varying Associations between Physical Activity and Affect Using Multilevel Modeling. Computational and Mathematical Methods in Medicine 2018, 1–11 (2018).

[32] Yang, Y. S., Ryu, G. W. & Choi, M. Methodological Strategies for Ecological Momentary Assessment to Evaluate Mood and Stress in Adult Patients Using Mobile Phones: Systematic Review. JMIR mHealth and uHealth 7, e11215 (2019).

[33] Scheffer, M. et al. A Dynamical Systems View of Psychiatric Disorders—Theory: A Review. JAMA Psychiatry 81, 618 (2024).

[34] Nelson, B., McGorry, P. D., Wichers, M., Wigman, J. T. W. & Hartmann, J. A. Moving From Static to Dynamic Models of the Onset of Mental Disorder: A Review. JAMA Psychiatry 74, 528 (2017).

[35] Epskamp, S. et al. Personalized Network Modeling in Psychopathology: The Importance of Contemporaneous and Temporal Connections. Clinical Psychological Science 6, 416–427 (2018).

[36] Rauschenberg, C. et al. Effects of AI4U training, a machine learning-based, adaptive ecological momentary intervention for personalized mental health promotion in youth: findings from a micro-randomized trial. Preprint at https://osf.io/mvyar (2024).

[37] Hiller, S. et al. Health-Promoting Effects and Everyday Experiences With a Mental Health App Using Ecological Momentary Assessments and AI-Based Ecological Momentary Interventions Among Young People: Qualitative Interview and Focus Group Study. JMIR mHealth and uHealth 13, e65106 (2025).

[38] Götzl, C. et al. Artificial intelligence-informed mobile mental health apps for young people: a mixed-methods approach on users’ and stakeholders’ perspectives. Child and Adolescent Psychiatry and Mental Health 16, 86 (2022).

[39] Brenner, M., Weber, E., Koppe, G. & Durstewitz, D. Learning Interpretable Hierarchical Dynamical Systems Models from Time Series Data. Preprint at https://arxiv.org/abs/2410.04814 (2024).

[40] Hess, F., Monfared, Z., Brenner, M. & Durstewitz, D. Generalized Teacher Forcing for Learning Chaotic Dynamics. In Proceedings of the 40th International Conference on Machine Learning, 13017–13049 (2023).

[41] Volkmann, E., Brändle, A., Durstewitz, D. & Koppe, G. A scalable generative model for dynamical system reconstruction from neuroimaging data. In NeurIPS Proceedings, Vol. 37, 80328–80362 (2024).

[42] Vaswani, A. et al. Attention is All you Need. In Advances in Neural Information Processing Systems, Vol. 30 (2017).

[43] Wu, N., Green, B., Ben, X. & O’Banion, S. Deep Transformer Models for Time Series Forecasting: The Influenza Prevalence Case. Preprint at http://arxiv.org/abs/2001.08317 (2020).

[44] Borsboom, D. & Cramer, A. O. Network Analysis: An Integrative Approach to the Structure of Psychopathology. Annual Review of Clinical Psychology 9, 91–121 (2013).

[45] Hofmann, S. G., Curtiss, J. & McNally, R. J. A Complex Network Perspective on Clinical Science. Perspectives on Psychological Science 11, 597–605 (2016).

[46] Hofmann, S. G. A Network Control Theory of Dynamic Systems Approach to Personalize Therapy. Behavior Therapy 56, 199–212 (2025).

[47] Brunton, S. L. & Kutz, J. N. Data-Driven Science and Engineering: Machine Learning, Dynamical Systems, and Control (Cambridge University Press, 2019).

[48] Cai, X. et al. State space model multiple imputation for missing data in non-stationary multivariate time series with application in digital Psychiatry. Preprint at https://arxiv.org/abs/2206.14343 (2022).

[49] Stone, A. A., Schneider, S. & Smyth, J. M. Evaluation of Pressing Issues in Ecological Momentary Assessment. Annual Review of Clinical Psychology 19, 107–131 (2023).

[50] Schumacher, L., Burger, J., Echterhoff, J. & Kriston, L. Methodological and Statistical Practices of Using Symptom Networks to Evaluate Mental Health Interventions: A Review and Reflections. Multivariate Behavioral Research 59, 663–676 (2024).

[51] Saeb, S. et al. Mobile Phone Sensor Correlates of Depressive Symptom Severity in Daily-Life Behavior: An Exploratory Study. Journal of Medical Internet Research 17, e175 (2015).

[52] Hojjatinia, S. et al. Dynamic models of stress-smoking responses based on high-frequency sensor data. npj Digital Medicine 4, 162 (2021).

[53] Epstein, D. H. et al. Prediction of stress and drug craving ninety minutes in the future with passively collected GPS data. npj Digital Medicine 3, 26 (2020).

[54] Rabbi, M., Klasnja, P., Choudhury, T., Tewari, A. & Murphy, S. Optimizing mHealth Interventions with a Bandit. In Digital Phenotyping and Mobile Sensing (eds Baumeister, H. & Montag, C.) 277–291 (Springer International Publishing, Cham, 2019).

[55] Elorrieta, F., Eyheramendy, S. & Palma, W. Discrete-time autoregressive model for unequally spaced time-series observations. Astronomy & Astrophysics 627, A120 (2019).

[56] Cunningham, J., Ghahramani, Z. & Rasmussen, C. Gaussian Processes for time-marked time-series data. In Proceedings of the Fifteenth International Conference on Artificial Intelligence and Statistics, 255–263 (2012).

[57] Koop, G., Pesaran, M. & Potter, S. M. Impulse response analysis in nonlinear multivariate models. Journal of Econometrics 74, 119–147 (1996).

[58] Rauschenberg, C. et al. Living lab AI4U - artificial intelligence for personalized digital mental health promotion and prevention in youth. European Journal of Public Health 31, ckab164.746 (2021).

[59] Öğretir, M., Ramchandran, S., Papatheodorou, D. & Lähdesmäki, H. A Variational Autoencoder for Heterogeneous Temporal and Longitudinal Data. In 21st IEEE International Conference on Machine Learning and Applications (ICMLA), 1522–1529 (2022).

[60] Brenner, M., Hess, F., Koppe, G. & Durstewitz, D. Integrating Multimodal Data for Joint Generative Modeling of Complex Dynamics. Preprint at http://arxiv.org/abs/2212.07892 (2024).

[61] Durstewitz, D., Koppe, G. & Thurm, M. I. Reconstructing computational system dynamics from neural data with recurrent neural networks. Nature Reviews Neuroscience 24, 693–710 (2023).

[62] Ansari, A. F. et al. Chronos: Learning the Language of Time Series. Preprint at http://arxiv.org/abs/2403.07815 (2024).

[63] Hewamalage, H., Ackermann, K. & Bergmeir, C. Forecast evaluation for data scientists: common pitfalls and best practices. Data Mining and Knowledge Discovery 37, 788–832 (2023).

[64] Izhikevich, E. M. Dynamical systems in neuroscience: the geometry of excitability and bursting (MIT Press, Cambridge MA, 2007).

[65] Kalman, R. E. A New Approach to Linear Filtering and Prediction Problems. Journal of Basic Engineering 82, 35–45 (1960).

[66] Bishop, C. M. Pattern recognition and machine learning (Springer, New York, 2006).

[67] Koppe, G., Toutounji, H., Kirsch, P., Lis, S. & Durstewitz, D. Identifying nonlinear dynamical systems via generative recurrent neural networks with applications to fMRI. PLOS Computational Biology 15, e1007263 (2019).

[68] Durstewitz, D. Advanced Data Analysis in Neuroscience (Springer International Publishing, Basel, 2017).

[69] Funahashi, K.-i. & Nakamura, Y. Approximation of dynamical systems by continuous time recurrent neural networks. Neural Networks 6, 801–806 (1993).

[70] Schmidt, D., Koppe, G., Monfared, Z., Beutelspacher, M. & Durstewitz, D. Identifying Nonlinear Dynamical Systems with Multiple Time Scales and Long-Range dependencies. In International Conference on Learning Representations, 29 (2021).

[71] Kimura, M. & Nakano, R. Learning dynamical systems by recurrent neural networks from orbits. Neural Networks 11, 1589–1599 (1998).

[72] Hanson, J. & Raginsky, M. Universal Simulation of Stable Dynamical Systems by Recurrent Neural Nets. In Proceedings of the 2nd Conference on Learning for Dynamics and Control, 384–392 (2020).

[73] Zhou, H. et al. Informer: Beyond Efficient Transformer for Long Sequence Time-Series Forecasting. Preprint at http://arxiv.org/abs/2012.07436 (2021).

[74] Grigsby, J., Wang, Z., Nguyen, N. & Qi, Y. Long-Range Transformers for Dynamic Spatiotemporal Forecasting. Preprint at http://arxiv.org/abs/2109.12218 (2023).

[75] Said, S. E. & Dickey, D. A. Testing for unit roots in autoregressive-moving average models of unknown order. Biometrika 71, 599–607 (1984).

